# Using data science to identify unusual treatment choices in England: illustrative findings of uncommon antipsychotics Pericyazine and Promazine

**DOI:** 10.1101/2022.03.22.22272504

**Authors:** Brian MacKenna, Helen J Curtis, Alex J Walker, Richard Croker, Orla Macdonald, Stephen JW Evans, Peter Inglesby, Dave Evans, Jessica Morley, Seb CJ Bacon, Ben Goldacre

## Abstract

**Background:** Data analysis can be used to identify signals suggestive of variation in treatment choice or clinical outcome. Analyses to date have generally focused on an hypothesis-driven approach.

**Methods:** Here we report an innovative hypothesis-blind approach (calculating chemical-class proportions for every chemical substance prescribed in each Clinical Commissioning Group and ranking chemicals by (a) their kurtosis and (b) a ratio between inter-centile differences) applied to England’s national prescribing data, and demonstrate how this identified unusual prescribing of two antipsychotics.

**Outcomes:** We identified that, while promazine and pericyazine are barely used by most clinicians, they make up a substantial proportion of all antipsychotic prescribing in two small geographic regions in England.

**Interpretation:** Data-driven approaches can be effective at identifying unusual clinical choices. More widespread adoption of such approaches, combined with clinician and decision-maker engagement could lead to better optimised patient care.

**Funding:** NIHR Biomedical Research Centre, Oxford; Health Foundation; National Institute for Health Research (NIHR) School of Primary Care Research and Research for Patient Benefit

**Research in context:** *Evidence before this study:* Identifying variation in clinical activity typically employs a traditional approach whereby measures are prospectively defined, and adherence then assessed in data. We are aware of no prior work using data science techniques hypothesis-blind to systematically identify outliers for any given treatment choice or clinical outcome (numerators) as a proportion of automatically generated denominators.

*Added value of this study:* Here we report an innovative hypothesis-blind approach applied to England’s national prescribing data, to identify chemical substances with substantial prescribing patterns between organisations. As illustrative examples we show that promazine and pericyazine, while rarely used by most clinicians, made up a substantial proportion of all antipsychotic prescribing in two small geographic regions in England.

*Implications of all the available evidence:* The choice of antipsychotics between English regions could be further investigated using qualitative methods to explore the implications for patient care. More broadly, data-driven approaches can be effective at identifying unusual clinical choices. More widespread adoption of such approaches, combined with clinician and decision-maker engagement could lead to better optimised patient care.

## Background

Since 2011 the NHS in England has openly shared detailed monthly general practice prescribing data at a high level of granularity, down to the level of individual doses, chemicals and brands, aggregated at individual general practice level. This data has supported original research on a broad range of topics as well as supporting a systematic audit and review programmes to support improvements in primary care prescribing.^1^

Our group produces OpenPrescribing.net, a free and widely used tool where anyone can explore the prescriptions dispensed at any practice in England and monitor prescribing patterns down to the level of individual brands, formulations and doses. OpenPrescribing offers data-driven feedback to assist regional and practice-level Medicines Optimisation teams and identify areas for review of which they may not otherwise have been aware. For example, we identify whether each NHS organisation is an outlier on more than 80 pre-defined measures covering a range of prescribing safety, cost-effectiveness and efficacy issues. We also calculate unique savings opportunities for each practice by making comparisons between brands/generics and formulations,^2^ and there is evidence that these savings are realised.^3^

Typically data science for service audit and quality improvement is hypothesis driven: identifying a targeted behaviour, and using data to measure achievement of that goal. Given the vast scale of NHS prescribing data (more than two billion rows of data covering 8,000 organisations during the past decade) and the vast range of clinical behaviours and potential signals for variation in care that may lie within this dataset, we set out to develop new hypothesis-blind data science techniques to identify new opportunities for service improvement driven by variation in care.

Our overall analytic aim was to prototype and describe methods to identify previously unknown signals of clinical interest in prescribing data. We ran a series of internal workshops to develop a short list of data science methods that might be used to identify prescribing behaviours that are unusually distributed across NHS organisations or regions. Here we briefly report the successful deployment of one such method (ranking chemicals by kurtosis and a ratio between inter-centile differences across all chemical-class pairs) and demonstrate how this identified high prescribing of unusual antipsychotics in two small regions of England.

## Methods

### Study design and data sources

We conducted a retrospective cohort study using open NHS prescribing data on all dispensed products prescribed by general practices in England, June-August 2017, extracted from the OpenPrescribing database. The dataset includes, for each practice, product and month of prescribing, the number of items prescribed (equivalent to the number of prescription forms on which each product appeared) and the total quantity (e.g. tablets, millilitres etc). Practices were grouped by their parent Clinical Commissioning Group (CCG), an NHS administrative region. England’s approximately 7000 NHS general practices were arranged into 207 CCGs in 2017.

### Data processing

All chemicals prescribed in England were assigned to a “class” of chemicals, using their British National Formulary (BNF) legacy code to identify the chemical’s relevant BNF sub-paragraph. We limited to chemicals in chapters 1-15 of the BNF (1511 prescribed chemicals), to exclude chapters not following a chemical/sub-paragraph structure, those which largely cover non-medicinal products such as dressings. For each chemical-class pair, we calculated the number of items (similar to a prescription in prescribing data) that was prescribed for each chemical as a proportion of the total items prescribed of all chemicals in its class (chemical-class proportion), for each CCG. To avoid including rarely prescribed classes of chemical that would generate spurious findings, we filtered to classes used by more than 50 CCGs, and excluded classes with the lowest total items prescribed (lowest two centiles).

### Ranking Chemical-Class Pairs by Kurtosis

Kurtosis is a numerical measure of the extent to which the tails of a given distribution are heavier or lighter than a normal distribution: overall, datasets with high kurtosis will tend to have more outliers than datasets with low kurtosis. Kurtosis is a good method for detecting an unknown number of outliers in a dataset.^4^ We calculated kurtosis for all chemical-class proportions by CCG, and ranked chemicals by this kurtosis (highest to lowest). We then additionally filtered to chemicals with a modal proportion of zero, i.e. those not prescribed at all by most CCGs (after rounding to three decimal places and filtering out CCGs with a zero denominator). We then alternatively ranked chemicals by a ratio calculated as the inter-centile range of the chemical-class proportion between the 95th-97th centiles (the top-prescribing CCGs) to the inter-centile range between 50th-95th centiles. Overall this approach sorts all chemicals into a ranked order where, for the most highly ranked chemicals, there are very substantial differences between CCGs in the extent to which that chemical is used in the context of all prescribing of all chemicals in its class. This ranking was used to prioritise chemical-class pairs for manual evaluation by clinical staff for signals of clinical interest.

For selected chemical-class pairs of clinical interest we generated a choropleth map using OpenPrescribing.net to visualise the geographical distribution of prescribing of each chemical as a proportion of its class. Data management was performed using Python and Google BigQuery, with analysis carried out using Python. Data, as well as all code for data management and analysis is openly available for inspection and re-use at https://github.com/ebmdatalab/kurtosis-pericyazine/tree/1.0.

## Results

We identified a range of chemicals exhibiting highly unusual patterns of usage across CCGs. Figure 1a shows the top 12 chemical substances identified through kurtosis ranking across all clinical areas. Of the chemicals with a high kurtosis, the two chemical substances with the highest ratio of 95th-97th:50-95th centile differences and with mode = 0 were two antipsychotics: promazine hydrochloride (ratio: 1·804, kurtosis: 43·61), and pericyazine (ratio: 0·880, kurtosis: 49·60) (figure 1b) and we present them here as an illustrative example. In the case of pericyazine (figure 2a), OpenPrescribing choropleth maps indicated prescribing was concentrated in the East of England, specifically around the Norfolk area, and promazine in North West England, particularly Bolton and the wider Greater Manchester area (figure 2b).

**Figure 1.**
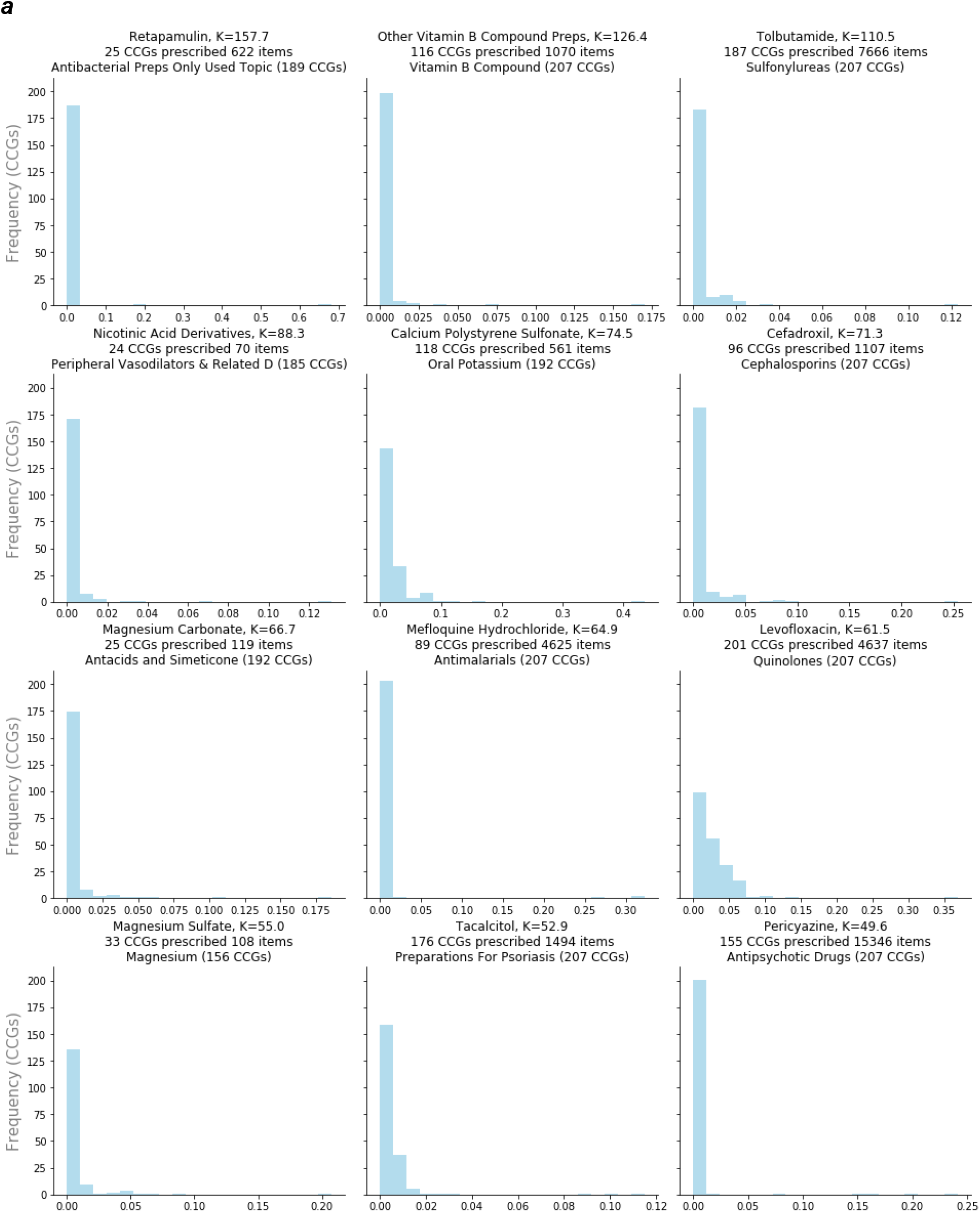
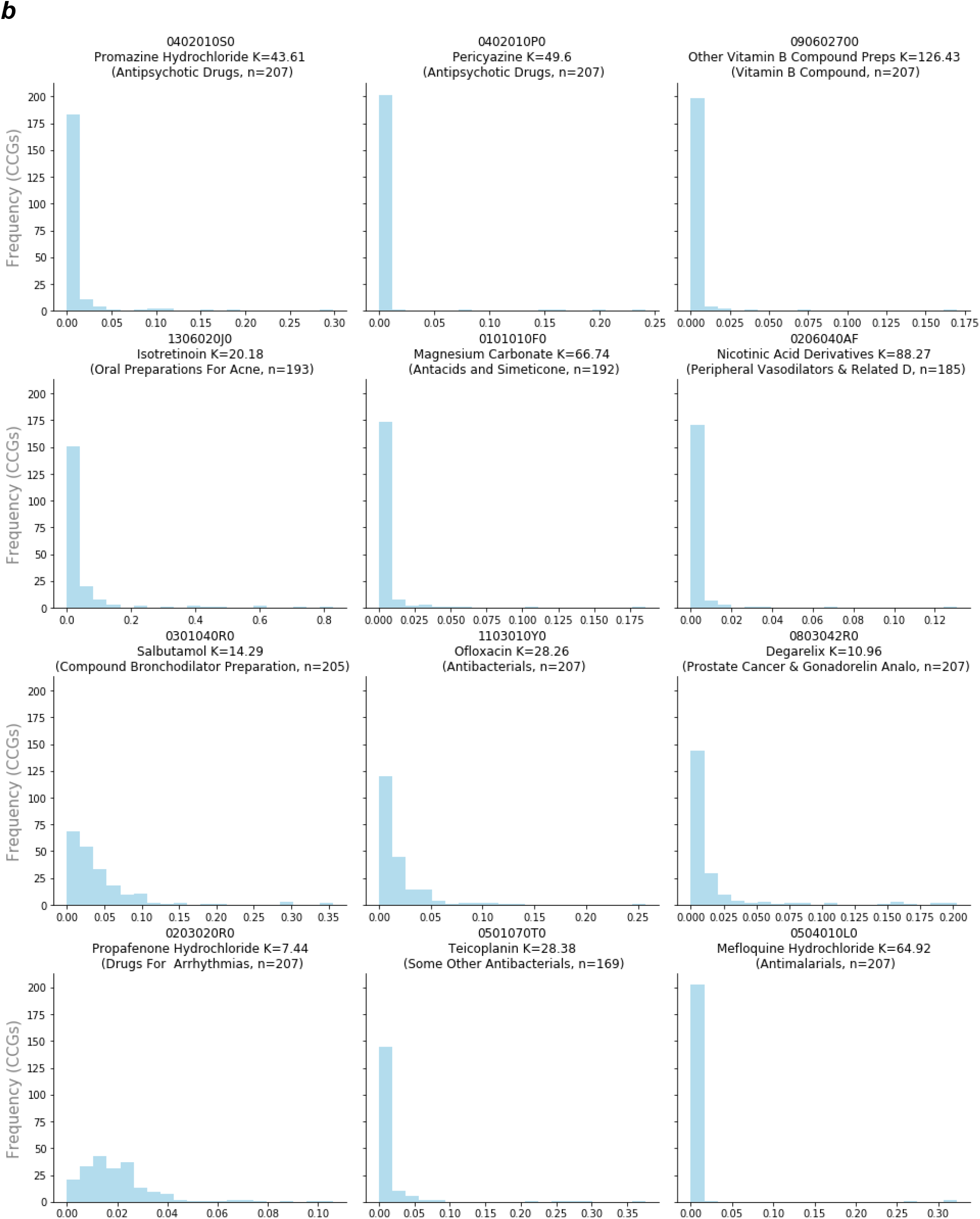
Chemical substances with unusual distributions of use across CCGs in England, June-August 2017: chemical-class proportions ranked using two alternative methods. Top 12 chemicals ranked by (a) kurtosis (K) of chemical-class proportions across CCGs and (b) ratio of 95th-97th:50-95th percentile differences of chemical-class proportions across CCGs (and filtered to those with mode=0). Chemical-class proportions were calculated as the number of items prescribed of each chemical as a proportion of items prescribed in its class, for each CCG. Each chart shows chemical-class proportion plotted against frequency of CCGs. The title indicates the number of CCGs (out of 207) which prescribed any of the chemical substance (and the class in parentheses), and how many items in total were prescribed in the study period.

**Figure 2:**
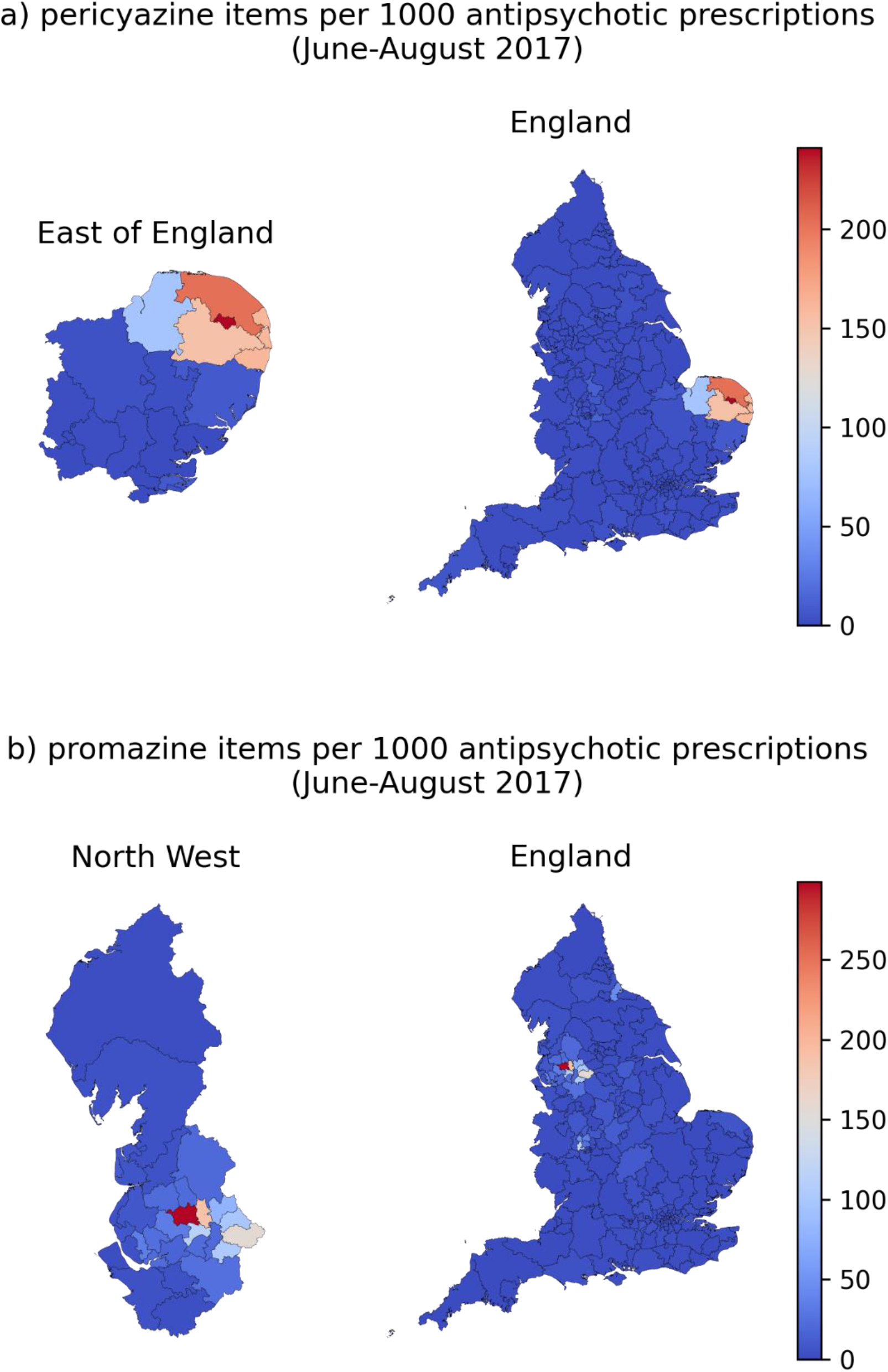
Total number of prescriptions for a) Pericyazine b) Promazine hydrochloride per 1000 antipsychotic prescriptions for all CCGs in England in June-August 2017.

## Discussion

Using new hypothesis-blind data science techniques for outlier detection we identified two unusual antipsychotic medications, in very limited use nationally, that are very commonly prescribed in two small geographic regions of England. Specifically, pericyazine makes up only 0·5% of all antipsychotic prescriptions nationally, but in Norfolk it represents 24% of all antipsychotic prescriptions; promazine hydrochloride 1·1% of all antipsychotic prescriptions nationally, however in Bolton it represents 30% of all antipsychotic prescriptions.

### Strengths and weaknesses

OpenPrescribing data includes all prescribing in all typical practices in England, thereby minimising the potential for obtaining a biased sample. The analysis was run on data covering Jun-Aug 2017; our work on reporting this data and investigating the rationale for this outlier was disrupted by the COVID-19 pandemic.

### Findings in Context

Pericyazine has been used infrequently for schizophrenia and for short-term adjunctive management of severe anxiety, psychomotor agitation, and violent or dangerously impulsive behaviour.^5^ There is no mention of pericyazine in any guideline on the National Institute for Health and Care Excellence (NICE) website^6^, the main source of clinical guidelines in England. A 2014 Cochrane review on pericyazine identified only five studies suitable for inclusion, could not determine pericyazine’s effect in schizophrenia given the low quality of evidence, and found a higher incidence of side effects compared to other antipsychotics^7^. A PubMed search identified only 73 publications that contain the word pericyazine in any way since 1965 compared with over 22,000 results for haloperidol, and 11,000 for risperidone.

Promazine is licensed in psychomotor agitation and agitation or restlessness in the elderly.^8^ It is not mentioned in any NICE guideline^9^, and appears in only 1,355 PubMed records (peaking in 1964). We are aware of no prior work using data science techniques hypothesis-blind to systematically identify outliers for any given treatment choice or clinical outcome in the manner outlined here.

### Policy Implications and Interpretation

We report only the fact of substantial deviation from national prescribing norms in these two small regions, and make no direct comment on the appropriateness of using these medications in any single patient, or in general. It was outside the scope of this data science project to engage in detailed qualitative or other work to understand the reasons for high usage of these two unusual antipsychotics in these two regions: however we note that promazine and pericyazine have previously appeared in treatment formularies for Greater Manchester and Norfolk respectively. In addition we note that antipsychotic medication is typically initiated in secondary care, with prescribing taken over in general practice.

The Department of Health and Social Care recently consulted on an ambitious plan to harness data to improve health delivery and outcomes^10^. The use of data to identify variation in clinical activity and outcomes is long established^11,12^ and recent flagship projects in the NHS such as RightCare and Getting it Right First Time (GIRFT) are focused on identifying and addressing variation in care. However these approaches typically rely on a traditional approach whereby desirable clinical activities or outcomes are prospectively defined by clinicians or commissioners, and adherence is then measured in the data. It is highly unlikely that these conventional methods would ever have identified the unusual prescribing behaviours reported in this paper. Similarly, it is likely that there are many further clinically interesting signals that could be identified by taking a variety of data-driven approaches to detecting unusual clinical activity or outcomes across the full universe of NHS data.

In our experience of running OpenPrescribing.net, the key barrier to better use of data for service improvement is an unhelpful cultural and practical divide between purely academic work on health data, and practical use of data in service analytics. This is exemplified by (in general) the use of different teams, different funding mechanisms, different institutions, and different data infrastructures. As the methods, data, and overarching objectives of both domains overlap substantially, we hope that funders and commissioners can help to bring these strands of work together.

### Future Research

Building on the methods described here we are now developing a range of interactive tools on OpenPrescribing.net to present candidate signals of interest for substantial divergence from national prescribing norms to individual practices, CCGs, and other key NHS organisational groupings such as Primary Care Networks and Integrated Care Systems. For this online service we expect to present a wide variety of signals at scale, without further context on evidence or guidelines, as a trigger for positive local discussion and further exploration by clinical or commissioning teams, and inviting feedback on whether they found the signals to be helpful in identifying any previously unrecognised opportunities to change local prescribing practices or understanding the reasons for any divergences.

### Conclusions

We describe a hypothesis-blind approach to identify candidates for audit and review in clinical practice, with examples highlighted of two very unusual antipsychotics used disproportionately in two small geographic areas of England.

## Data Availability

All data produced are available online at https://github.com/ebmdatalab/kurtosis-pericyazine

## Administrative

### Funding

This work was supported by The NIHR Biomedical Research Centre, Oxford, A Health Foundation grant (Award Reference Number 7599); A National Institute for Health Research (NIHR) School of Primary Care Research (SPCR) grant (Award Reference Number 327); the National Institute for Health Research (NIHR) under its Research for Patient Benefit (RfPB) Programme (Grant Reference Number PB-PG-0418-20036) and by the National Institute for Health Research Applied Research Collaboration Oxford and Thames Valley. The views expressed in this publication are those of the author(s) and not necessarily those of the NIHR, NHS England or the Department of Health and Social Care. Funders had no role in the study design, collection, analysis, and interpretation of data; in the writing of the report; and in the decision to submit the article for publication.

## Ethical approval

This study uses exclusively open, publicly available data, therefore no ethical approval was required.

## Guarantor

BG is guarantor.

## Contributorship

BG conceived the study with input from SE. HJC, AJW designed the methods. HJC collected and analysed the data with methodological and interpretation input from AJW, BMK, RC & BG. HJC, AJW and SB all directly accessed and verified the underlying data, as published by NHSBSA. All authors confirm that they had full access to all the data in the study and accept responsibility to submit for publication. BMK drafted the manuscript with input from RC, HJC, SE, BG, OM & JM. All authors contributed to and approved the final manuscript.

SB was lead engineer on the associated website resource with input from DE & PI. BG supervised the project and is guarantor.

## Data sharing

This study uses exclusively open, publicly available data. Processed data is additionally available within the study repository along with all analysis code: https://github.com/ebmdatalab/kurtosis-pericyazine/tree/1.0.

